# Cuff Algometry Induces Large Yet Variable Conditioned Pain Modulation Effects

**DOI:** 10.1101/2025.09.29.25336881

**Authors:** Joseph L. Taylor, Timothy Lawn, Olivia S. Kowalczyk, Thomas Graven-Nielsen, Matthew A. Howard, Kirsty Bannister

**Author notes:** **Corresponding author:** Joseph L. Taylor, Address: L1.08, 16 De Crespigny Park, Centre for Neuroimaging Sciences, Institute of Psychiatry, Psychology and Neuroscience, King’s College London, London, SE5 8AF. These authors contributed equally joint first (Joseph L. Taylor, Timothy Lawn). joint last authorship (Matthew A. Howard, Kirsty Bannister). **Other author emails:**.

## Abstract

Conditioned pain modulation (CPM) paradigms provide a proxy measure of activity in the descending pain modulatory system. Cuff-pressure-algometry offers a standardised CPM assessment tool although comprehensive validation in large samples is lacking. To address this, we pooled cuff-algometry CPM data from 324 healthy participants across 8 studies. CPM magnitude was calculated as pain detection (PDT) and tolerance (PTT) threshold changes, assessed on the dominant leg in the presence and absence of a painful “conditioning” cuff stimulus on the contralateral leg. CPM-effects were robust for both changes in PDT and PTT (p<0.001). Using a classification approach where a ≥20% change in threshold designated a CPM responder, 69% of participants were CPM-responders for PDT and 59% for PTT. Test-retest reliability data were assessed in a subset of participants (n=72; interval 16.49±18.39days) using intraclass correlation coefficients (ICC). Test-retest reliability was *poor* for CPM-effects (ICC=0.25-0.37) despite *moderate-to-good* reliability for PDT and PTT (ICC=0.69-0.87). Responder classification showed *none*-to-*minimal* agreement across sessions (Cohen’s κ=0.17-0.21), with 38% of participants switching classification for both PDT and PTT. Bootstrap analysis revealed that smaller samples provide highly variable ICC estimates, potentially explaining discrepancies with previous reliability reports. Despite producing large group-level CPM-effects, *poor* test-retest reliability of cuff algometry suggests it captures dynamic, state-dependent processes rather than a stable trait-like individual characteristic. This highlights the need to consider the temporal instability of CPM when interpreting data and considering its deployment within precision pain medicine.

## Introduction

Conditioned pain modulation (CPM) is the behavioural phenomenon whereby an individual’s perception of a noxious “test” stimulus is modulated by concurrent application of a second noxious “conditioning” stimulus. Psychophysical CPM paradigms are proposed to indicate efficacy of descending pain modulatory circuits [38], with dysfunction reported in several chronic pain conditions [33,46]. Despite initial promise as a biomarker [25], CPM does not consistently correlate with patients’ pain intensities nor duration, and while many studies report case-control differences, clinical utility remains elusive [7]. A recent study reported the impact of varying the conditioning stimulus timing on CPM’s ‘sensitivity’, highlighting the impact of methodological differences on CPM functionality as a pain-related biomarker [11]. Despite calls for standardisation [48], substantial methodological variability in stimulus timing, modality, and intensity between studies continues to limit the utility of CPM as a biomarker for chronic pain [7].

Cuff-algometry is a contemporary stimulus modality for CPM paradigms and a strong candidate for standardised testing. It involves using tourniquet cuffs (typically placed around the calf muscles) to apply ramps of gradually increasing pressure stimulation to derive pain detection and pain tolerance thresholds for each leg. Following this, a static pressure stimulus is applied to one leg, to serve as a noxious conditioning stimulus, whilst simultaneously thresholds are re-assessed at the other leg. This paradigm allows the conditioning stimulus intensity to be personalised, facilitating standardisation of perceived painfulness across individuals. The procedure is methodologically simple, fast, computer-controlled and largely user-independent, providing a balance of scalability with standardisation and reproducibility of application.

Initial clinical work has shown that cuff-algometry CPM assessment is sensitive to both differences between patient groups [43,44] and case-control comparisons [34], and may also predict post-surgical pain outcomes [32]. Several psychophysical aspects of this paradigm have already been characterised, including changes in thresholds due to repeated application [16,35], impacts of cuff location and stimulus intensity [12,41], and responses to sensitisation and analgesia [36]. Initial assessments have shown *good*-to-*excellent* test-retest reliability [12], comparable to other stimulus modalities [18,45]. However, these assessments used only modest sample sizes, with little consensus on defining a “functional” CPM response and wide variation in classification thresholds [5,34,43]. Comprehensive characterisation in a large cohort of healthy individuals is a requisite step towards validating the clinical potential of CPM. To date, such examination is lacking.

In this work, we pooled cuff-pressure CPM assessments from eight studies with identical psychophysical methodologies. We perform a large-scale characterisation of the protocol, considering both single-session (n=324) and test-retest (n=72) designs. Our primary aims were to investigate whether cuff-algometry CPM induces robust group-level effects and to see whether these are reliable across sessions, both in terms of absolute values and consistency of binary responder/non-responder classification. Additionally, we examined the relationships between baseline pain thresholds and the recorded CPM effects.

## Methods

### Source data

Data from 324 individuals were pooled from eight research studies performed on separate campuses at King’s College London. In two of the studies the protocol was repeated twice in identical, separate sessions, creating a test-retest sub-sample of 72 individuals. Data from two of the contributing studies have been published [8,31]. Ethical clearance for this (ID: LRS-22/23-36682) and all contributing studies was granted by the King’s College Health Research Ethics Committee. All studies were conducted in accordance with the revised Declaration of Helsinki. Consent for data to be used in future research studies was given by all participants.

All studies recruited participants aged 18 years or older, with no ongoing pain, no ongoing cardiovascular, neurological or pain medication use, no pregnancy, no diagnosed mental health conditions, and no central nervous system disorders. In addition to the CPM data, we recorded age, sex, and dominant leg laterality. Study-specific characteristics and any methodological differences are summarised in Table 1.

**Table 1.** Study Specific Demographics and Methods. DFNS QST: German Research Network on Neuropathic Pain Quantitative Sensory Testing Protocol, WUR: Wind-up Ration Test, MRI: Magnetic Resonance Imaging

| Study (Bold studies contributed to test-retest sample) | N | Number of Experimenters | Sex (M/F/Missing) | Age in Years Mean (SD) | Additional Inclusion/Exclusion Criteria | Additional Screening | Reimbursement | Tasks Completed Before Cuff Tests |
| --- | --- | --- | --- | --- | --- | --- | --- | --- |
| 1 | 35 | 1 | 17/18/0 | 23.8 (2.6) | No more than 5 cigarettes or 6 caffeinated drinks per day | Drug and alcohol screening, MRI contraindications | £23 per hour | DFNS QST |
| 2 | 32 | 2 | 21/11/0 | 25.5 (5.9) | No more than 5 cigarettes or 6 caffeinated drinks per day | Drug and alcohol screening, MRI contraindications. | £23 per hour | Drug screening, Sensory Testing familiarisation and thresholding, autonomic measurements, psychometry |
| 3 | 11 | 2 | 4/7/0 | 28.6 (2.6) | None | None | None | Heat and pressure CPM testing |
| 4 | 45 | 3 | 17/28/0 | 32.8 (11.8) | None | None | £10 | DFNS QST |
| **5** | 67 | 1 | 14/53/0 | 25.1 (7.9) | None | None | £50 | None |
| 6 | 40 | 2 | 10/30/0 | 27.9 (9.1) | None | None | £25 | DFNS QST WUR tests |
| **7** | 39 | 4 | 13/25/1 | 29.3 (10.2) | None | None | None | DFNS QST |
| 8 | 55 | 1 | 23/32/0 | 24.2 (5.8) | No more than 5 cigarettes or 6 caffeinated drinks per day. | Drug and alcohol screening, MRI contraindications. Self-harm Inventory score less than 5. | £23 per hour | DFNS QST |

### Pain detection threshold and pain tolerance threshold

Participants undertook a protocol incorporating a standardised cuff CPM paradigm, as previously described [4,5,12,13,16]. In brief, participants had a tourniquet cuff (VBM Medizintechnik GmbH, REF: 20-54-522) attached to each calf, with inflation controlled using the cuff pressure algometry system (Nocitech CPAR, Inventors’ Way ApS, Denmark). Pain thresholds were assessed using pressure ramps inflated at 1 kPa/s. The first ramp was applied to the dominant leg (Figure 1A), followed by the non-dominant leg (Figure 1B). Participants used an electronic 10 cm long visual analogue scale (VAS) anchored at “no pain” (0 cm) and “worst pain imaginable” (10 cm) to rate their perceived pain. When participants could no longer tolerate any more pain, they pressed a button to stop inflation.

**Figure 1.**
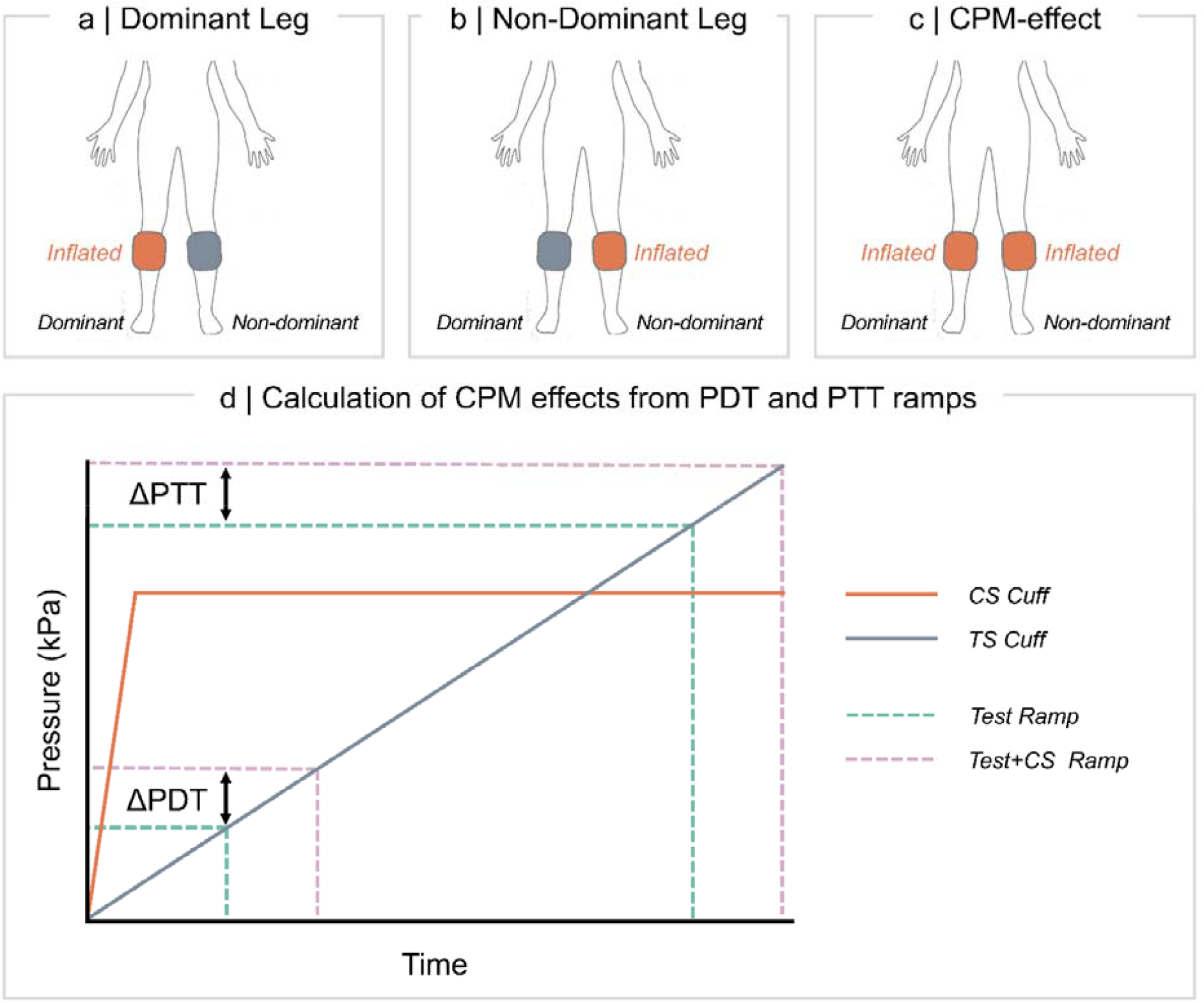
Psychophysics overview. Configuration of cuffs for assessment of PDT and PTT on the **(a)** Dominant and **(b)** Non-Dominant legs followed by reassessment of thresholds on the dominant leg in the presence of conditioning **(c). (d)** During each ramp, pressure increases with 1 kPa/s. PDT is defined as the pressure at which stimulation becomes painful (> 1 cm on the VAS), and PTT as the maximum tolerated pressure. CPM effects are computed as the difference (delta) in PDT and PTT, respectively, between assessment with conditioning **(c)** and without **(a)** on the dominant leg.

Each pressure ramp provided two psychophysical outputs. Pain Detection Threshold (PDT) was defined as the cuff pressure at which participants first moved the VAS slider away from the “no pain” anchor (instrumentalised as 0.1 cm on the VAS). Pain Tolerance Threshold (PTT) was defined as the maximum pressure (kPa) participants could tolerate before pressing the stop button.

All ramps were safety-limited at 97 kPa, after which cuffs automatically deflated to prevent injury. If so, PTT could not be accurately recorded and that participant was not used for further PTT analysis. Leg dominance was assessed by self-report and additionally prompted by asking participants with which leg they would kick a football [27].

### Conditioned pain modulation

CPM was assessed using concurrent cuff inflation as the conditioning stimulus (CS, Figure 1C). The CS cuff on the non-dominant leg was swiftly inflated to a static pressure equivalent to 70% of the PTT recorded on the non-dominant leg [47]. Once the CS pressure was reached and maintained, the test stimulus (TS) cuff on the dominant leg began inflating at 1 kPa/s, using an identical ramp protocol to the baseline measurements. Participants received the same VAS rating instructions as during baseline measurements, but were specifically instructed to rate only the painfulness of the TS on the dominant leg and to ignore the pressure applied to the non-dominant leg during the CPM assessment.

CPM magnitude was calculated as the difference in PDT and PTT, respectively, recorded during conditioning and at baseline (e.g. conditioned PDT minus baseline PDT). Thus, positive CPM-effects indicate increased pain thresholds (a hypoalgesic effect) in the presence of the conditioning stimulus.

### Classifying CPM responders and non-responders

Participants were classified as CPM responders or non-responders based on the magnitude of their pain threshold changes. Specifically, responders were designated as those showing ≥20% increase in both PDT and PTT thresholds during conditioning, a criterion previously employed in patient populations [43,44]. The tradition of applying a classification threshold to PDT and PTT changes, rather than binarizing around a change of 0, is essential to account for the measurement error inherent in repeating a test stimulus. However, these measurement error thresholds require test-retest data and can only be generalised out-of-sample to comparable cohorts. The 20% change criterion can be applied without requiring test-retest in the same participants and allows some direct comparison against patient populations. Participants who had sufficiently high PTT thresholds such that they could not achieve a 20% increase due to the safety limit were excluded from PTT classification analyses.

### Statistical analysis

Data are presented as mean values and standard deviation. All statistical analyses were conducted using R version 4.4.1. Group-level CPM-effects were assessed using linear mixed-effects models (lmer function from lme4 package [2,23]), with participant ID defined as a random intercept to account for repeated measures. Models included fixed effects for condition (e.g. PDT vs. PDT with conditioning), age, sex, and study. Separate models were fitted for PDT and PTT outcomes. Whilst sex differences were not the main focus of this work, we report mixed effects models examining the interaction between condition and sex within Supplementary Figure 1. We computed *p*-values for fixed effects via Satterthwaite approximation. The significance level was set at α=.05 for all analyses

The main CPM models took the following form:

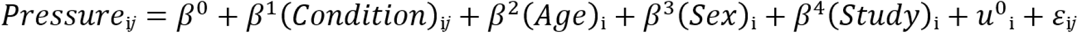

Where Pressure = PDT or PTT, Condition = baseline or conditioning, i = participants, j = conditions (baseline/conditioning), u_0i_ = the random intercept for participant i, and ε_ij_ = the residual error term.

Exploratory interrelationships between psychophysical measures were examined using linear models also accounting for age, sex, and study as covariates. These analyses investigated: (1) the relationship between conditioning pressure intensity and CPM-effect, (2) associations between baseline pain thresholds and CPM-effects, and (3) concordance between dominant and non-dominant leg measurements.

Test-retest reliability (n = 72) was assessed using multiple metrics. Intraclass Correlation Coefficients (ICC) were calculated using the two-way mixed-effects model for absolute agreement [ICC(2,1)] from the irr package [10]. ICCs were interpreted according to the following criteria: <0.50 *poor*, 0.50-0.75 *moderate*, 0.76-0.90 *good*, >0.90 *excellent* reliability [20]. We additionally report Pearson Correlation Coefficients, Standard Error of Measurement (SEM), and Coefficient of Variation (CoV). To examine the effect of sample size on reliability estimates, bootstrap analysis simulated ICC values across sample sizes from 10 to the full dataset (increments of 5). For each target sample size, we created computed ICC(2,1) values for 1000 bootstrap samples utilising replacement. Median ICC and 95% confidence intervals (2.5th-97.5th percentiles) summarized the bootstrap distributions. Consistency of responder/non-responder classification across sessions was assessed using Cohen’s kappa (<0.20 *none*, 0.21-0.39 *minimal*, 0.40-0.59 *weak*, 0.60-0.79 *moderate*, >0.80-0.90 *strong*, > 0.90 *almost perfect* [26]).

## Results

### Participants, data quality and ceiling effects

The final sample had a mean age of 26.9 years (SD = 8.53, 32 missing values) and comprised 119 male and 204 female participants (1 missing value). Detailed information regarding missing values is presented in Supplementary Table 1.

Analyses were conducted on 311 participants for PDT analyses and 257 for PTT analyses. This follows list-wise exclusion of all participants with missing sex or age data, in addition to 56 participants (17.28%) being excluded from PTT analyses for reaching the safety threshold. For responder classification analyses, a separate 53 participants (16.36%) were excluded because their baseline PTT was sufficiently high that a 20% increase would have surpassed the algometer’s safety limit.

The test-retest subsample comprised 72 participants (mean age = 26.3 years, SD = 8.1; 17 males, 55 females) with a mean inter-session interval of 16.5 days (SD = 18.4). Participants were excluded from PTT analyses if they exceeded the safety-limit in at least one session, resulting in sample sizes of 56 for baseline PTT (22.22% excluded), 49 for PTT during conditioning (31.94% excluded), and 48 for the PTT CPM-effect analyses (33.33% excluded). A separate 25 participants (34.72%) were excluded from PTT responder classification analyses as their baseline thresholds were too high to permit a 20% increase without exceeding the safety limit. There were no missing data exclusions in the subsample.

### Group-level CPM effect

PDTs increased from baseline (M = 21.86 kPa, SD = 10.05) to conditioning conditions (M = 30.78 kPa, SD = 15.57, *b* = 8.90, *t*(310) = 15.30, p < .001; Figure 2a). Similarly, PTTs increased from baseline (M = 47.48 kPa, SD = 17.45) to conditioning (M = 57.72 kPa, SD = 19.54, *b* = 10.24, *t*(256) = 21.74, p < .001; Figure 2b). The mean PDT CPM-effect was 8.90 kPa (SD = 10.26, 95% CI [7.76, 10.04]) and mean PTT CPM-effect was 10.24 kPa (SD = 7.55, 95% CI [9.40, 11.09]). Those with a greater PDT CPM-effect also showed a higher effect for PTT (*b* = 0.24, *t*(245) = 4.55, p < .001; Figure 2e). Using the 20% threshold, fewer participants qualified as CPM responders for PTT (59%) than PDT (69%). Despite the significant correlation between measures, only 36% of participants qualified as CPM responders on both PDT and PTT (Figure 2f). The PDT and PTT were higher in males compared with females, but no significant sex effects were found for PDT and PTT CPM-effects (Supplementary Figure 1.).

**Figure 2.**
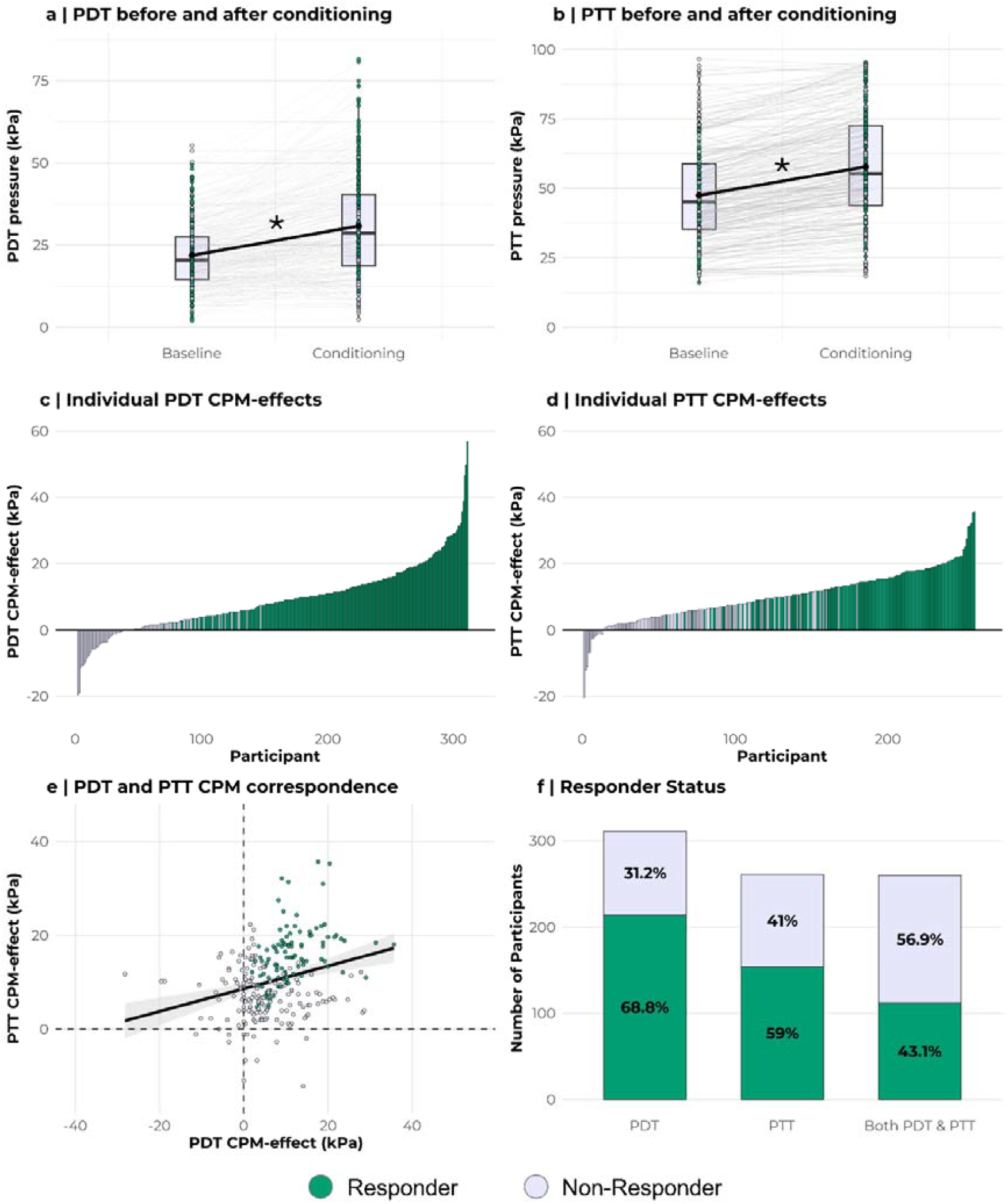
Group level CPM effects. **(a)** Pain Detection Thresholds (PDTs) and **(b)** Pain Tolerance Thresholds (PTTs) measured before and during the conditioning stimulus. **(c)** PDT CPM-effect and **(d)** PTT CPM-effect for each participant sorted by magnitude. **(e)** Correlation between PDT and PTT CPM-effects. **(f)** Percentage of sample classified as responders for PDT and PTT, together with coincidence of the two.

### Interrelationships between psychophysical measures

Greater conditioning pressure was associated with a larger increase in thresholds for both the PDT (*b* = 0.64, *t*(299) = 8.44, *p* < .001, Figure 3a) and PTT CPM-effects (*b* = 0.27, *t*(245) = 3.74, *p* < .001, Figure 3d). A higher baseline PDT threshold was associated with a greater increase in thresholds in the presence of the CS (*b* = 0.16, t(300) = 2.65, p = .009, Figure 3b). This however was not true for baseline PTT (*b* = 0.05, *t*(246) = 1.78, *p* = .0762, Figure 3e). Finally, there was strong concordance between thresholds on the dominant and non-dominant legs for PDT thresholds (*b* = 0.74, *t*(305) = 17.5, *p* < .001, Figure 3c) and PTT thresholds (*b* = 0.85, *t*(246) = 23.6, *p* < .001, Figure 3f). Overall, there was a positive manifold across all the thresholds measured, indicating participants tended to show higher or lower thresholds across all measurements in general (Supplementary Table 2).

**Figure 3.**
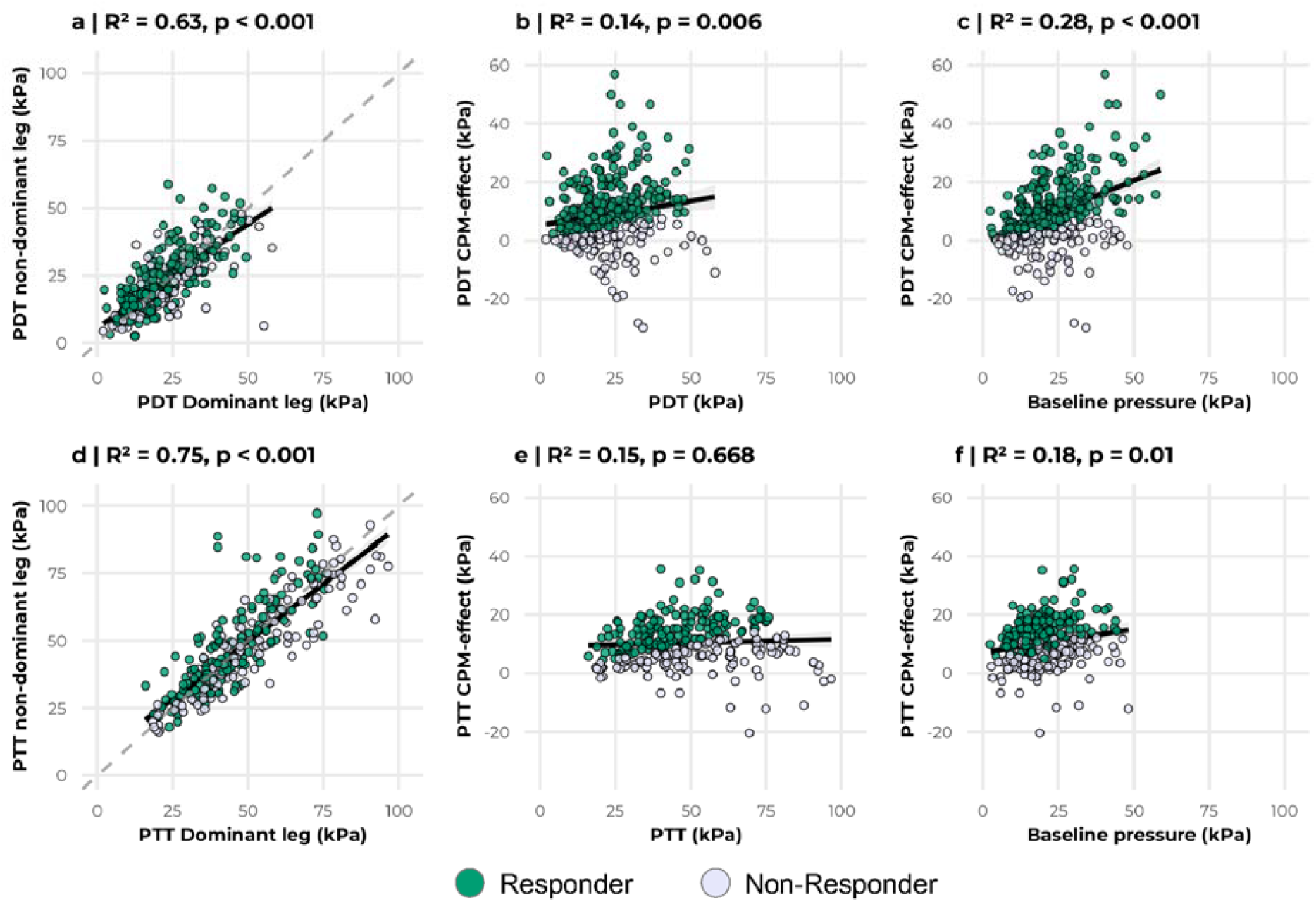
Psychophysical interrelationships. Correlation between the conditioning pressure and CPM effect, between the baseline threshold and the CPM effect, and between the dominant and non-dominant leg for PDT/PDT CPM-effect(**(a), (b)** and **(c)** respectively) and for PTT/PTT CPM-effect (**(d), (e)** and **(f)** respectively).

### Test-retest reliability

Reliability patterns differed markedly between raw thresholds and CPM effects. Individual PDT and PTT measurements demonstrated *moderate-to-good* test-retest reliability, with strong correlations and low measurement error. In contrast, PDT and PTT CPM-effects showed *poor* reliability, with weak correlations, high coefficients of variation, and poor ICCs (Table 2). Considering the CPM-effect as a relative effect (percentage change from baseline) rather than an absolute effect also demonstrated *poor* reliability between sessions (Supplementary Figure 2.)

**Table 2.**
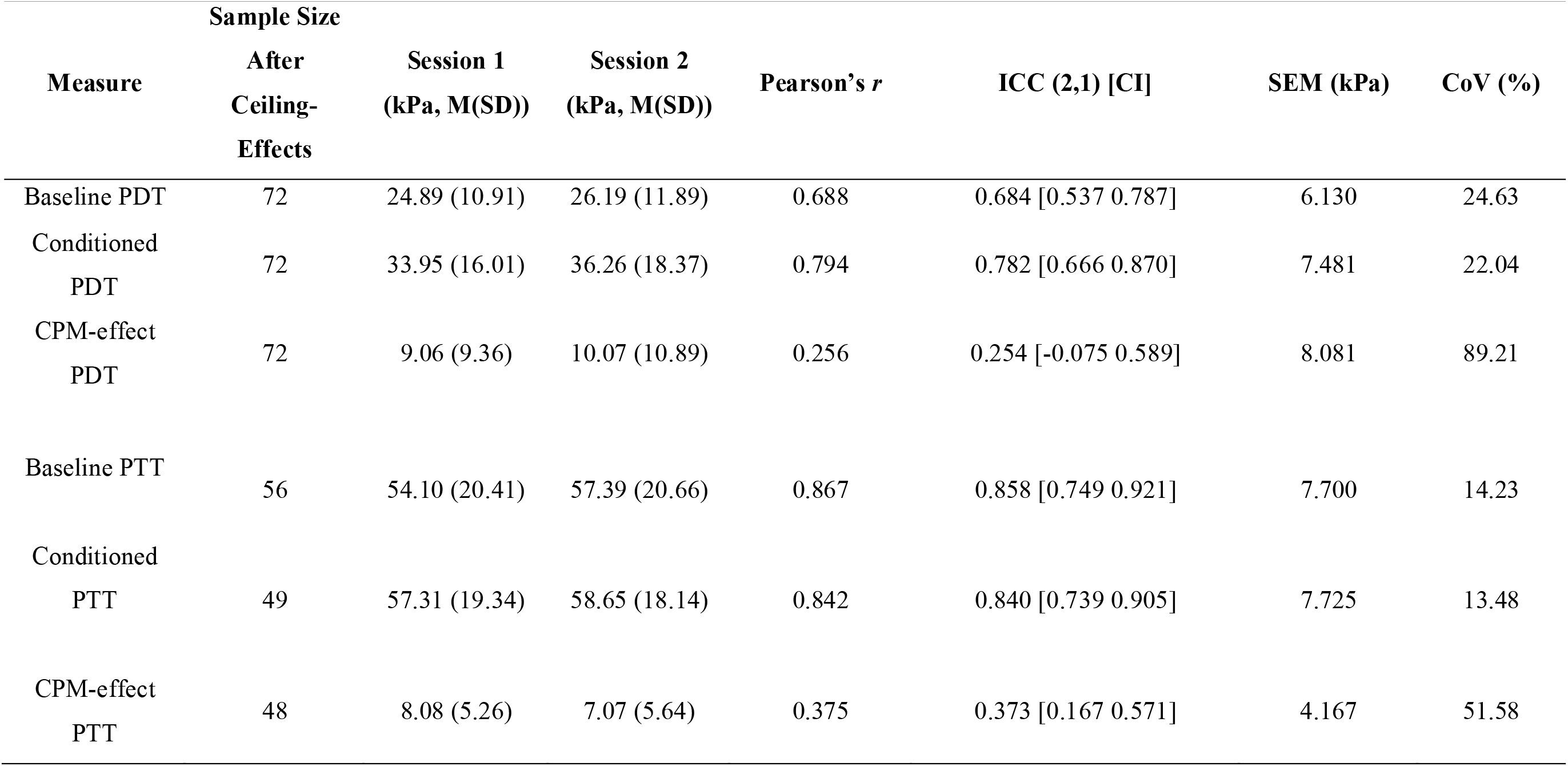
Descriptive and Reliability Statistics for the Test-Retest Sample. ICC: Intra-class Correlation Coefficient, SEM: Standard Error of Measurement, CoV: Coefficient of Variation, PDT: Pain Detection Threshold, PTT: Pain Tolerance Threshold

Given the large variability in CPM responses (Figure 2), we examined the effect of sample size on ICC estimates using bootstrap analysis. For the PDT CPM-effect, median ICC decreased from 0.314 (95% CI [-0.327, 0.703]) at n=25 to 0.268 (95% CI [-0.092, 0.580]) at our full sample (n=72), with substantial reduction in confidence interval width (Figure 4a). For the PTT CPM-effect, ICCs remained more stable across sample sizes: 0.365 (95% CI [0.029, 0.648]) at n = 25 versus 0.372 (95% CI [0.163, 0.566]) at full sample size (n = 48; Figure 4b). However, a similar widening of confidence intervals was observed with decreasing sample size.

**Figure 4.**
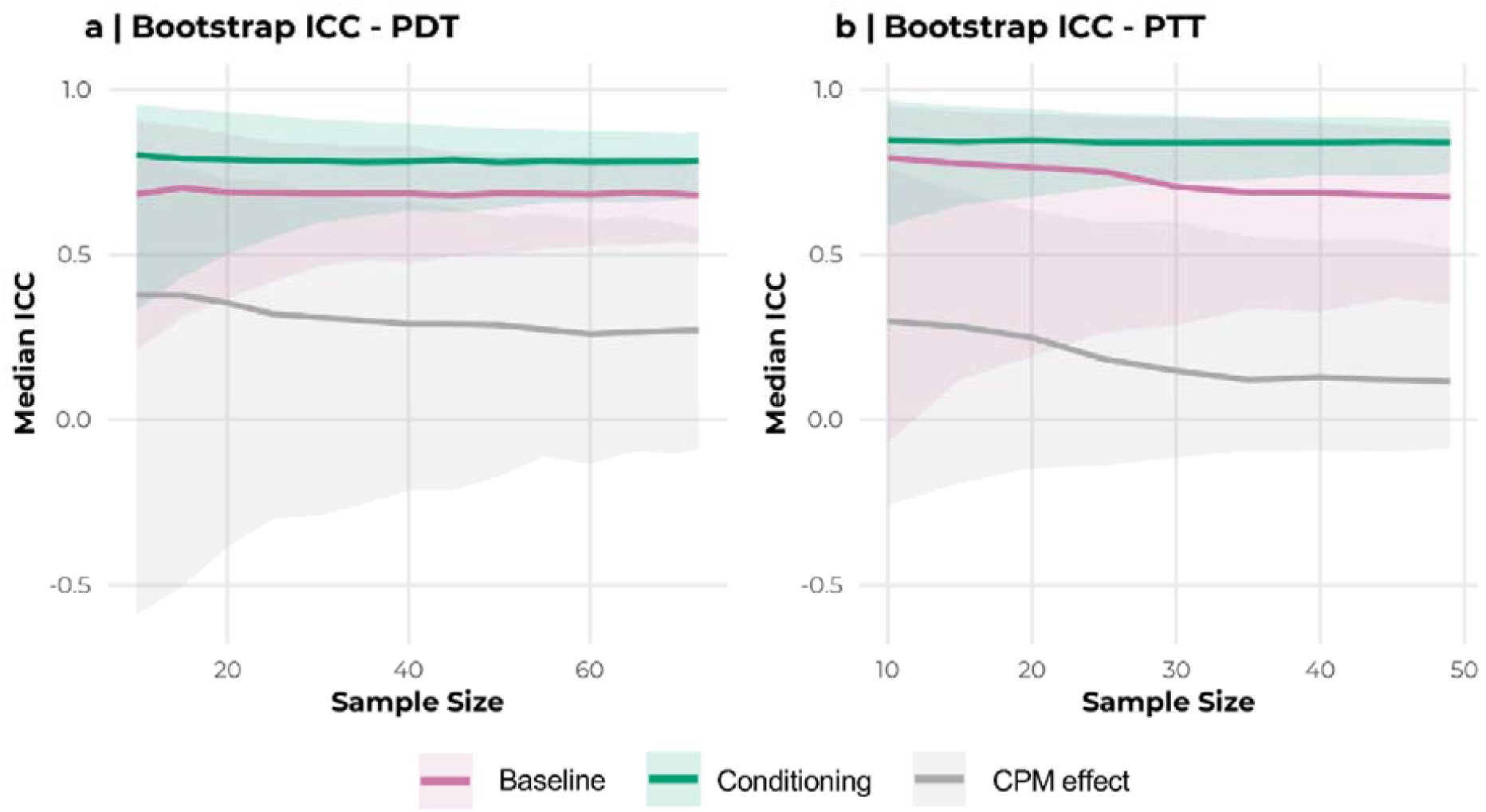
The effect of simulated sample size on reliability. Median ICCs taken from 1000 bootstrapped samples with replacement across a range of sample sizes for **(a)** PDT and PDT CPM-effect measurements and **(b)** PTT and PTT CPM-effect measurements. Shaded area represents the 95% confidence interval.

### Between session changes in responder/non-responder status

Responder classification showed *none-*to-*minimal* agreement across sessions (Figure 5). For PDT (n = 72), 50 participants were classified as responders in session 1 and 45 in session 2, with 27 participants (37.50%) switching classification. Specifically, 16 lost and 11 gained responder status (Cohen’s κ = 0.17; Figure 5a). For PTT (n = 45 after ceiling exclusions), 20 were responders in session 1, and 13 in session 2, with 17 participants (37.78%) switching classification. Specifically, 12 lost and 5 gained responder status (Cohen’s κ = 0.21; Figure 5b). Classification changes showed minimal concordance between PDT and PTT measures, with only 4 of 12 who lost PTT responder status also losing PDT responder status. Similarly, only 1 of 5 new PTT responders also gained PDT responder status. Whilst responder rates in the test-retest subsample for PDT match closely to that of the larger main sample, PTT responder rates were distinctly lower at 44/28% compared to 59% in the full dataset. The choice of threshold did not substantially alter Cohen’s Kappa values, with comparably poor reliability across a range of thresholds from 10-30% (Supplementary Figure 3).

**Figure 5.**
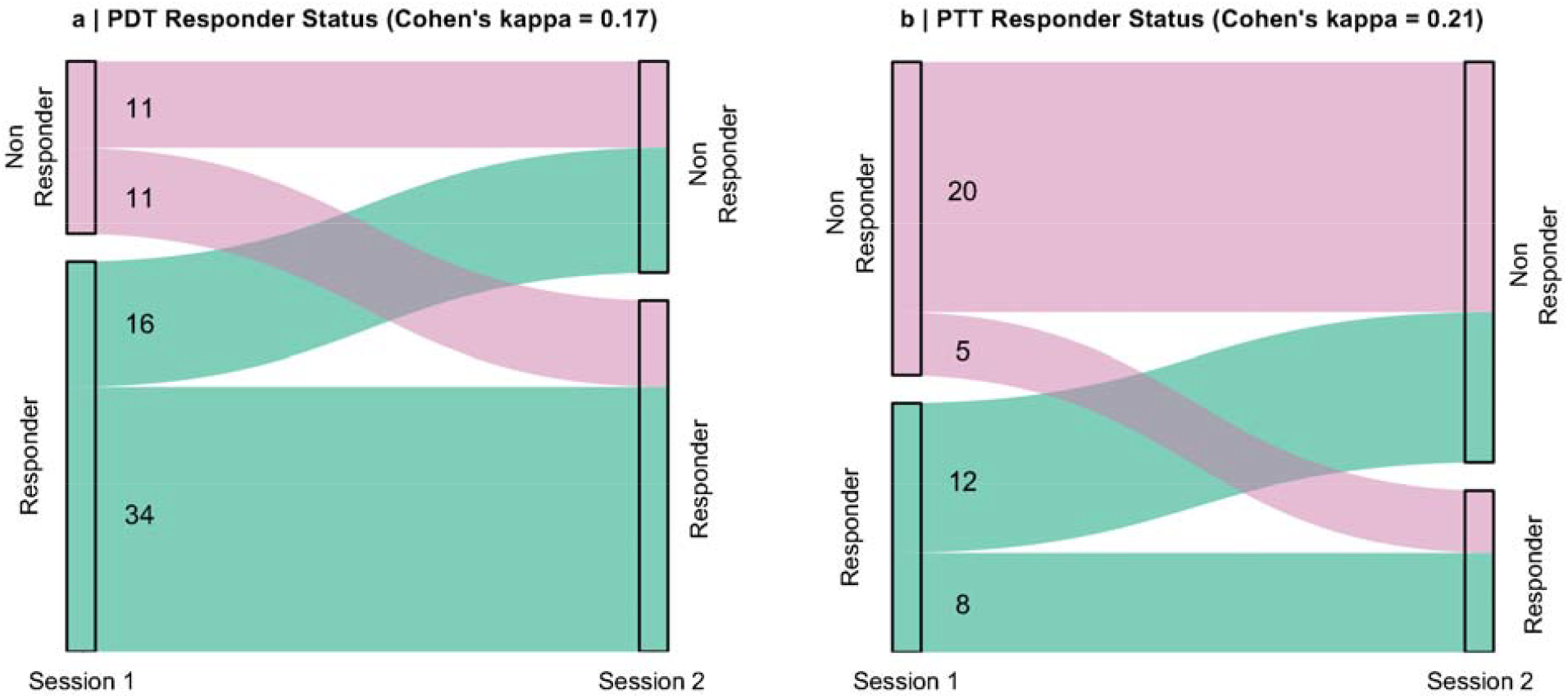
Responder classification stability across sessions. Transitions in responder status between sessions for **(a)** PDT (Cohen’s κ = 0.17) and **(b)** PTT (Cohen’s κ = 0.21). Width of flows represents number of participants.

## Discussion

This analysis provides a comprehensive examination of the CPM-effect upon application of a standardised cuff algometer paradigm in a large healthy cohort. We demonstrated robust group-level CPM-effects for both PDT and PTT, echoing prior accounts. By contrast, test-retest reliability of CPM-effect magnitudes and responder classification were poor. We propose that CPM-effects capture a dynamic, state-dependent process rather than a stable trait characteristic. Here we discuss both biological and methodological factors that may underpin this poor reliability.

Within a single session, cuff-pressure-algometry CPM demonstrated a strong group effect, with marked increases in the magnitude of both PDT and PTT observed in the presence of painful contralateral conditioning. The magnitude of these effects accords with previous accounts, with near identical estimates for PDT CPM-effects in studies comprising large (N > 60) samples [34]. We interpret prior reports of both larger and smaller magnitudes of CPM-effect simply in relation to increased variability expected in smaller samples, often featuring only 20 individuals or fewer [4,5,16]. There are no existing large sample estimates for PTT CPM-effects, but reports from multiple smaller studies suggest they vary even more than for PDT CPM-effects [4,5,16].

Approximately 67% of our participants were designated as PDT CPM responders. Our chosen responder classification threshold has not been previously imposed in healthy individuals using cuff algometry. However, investigations in mixed chronic pain populations have shown lower responder rates of approximately 50% [43,44], broadly supporting hypotheses of dysfunctional CPM responses in chronic-pain patients and a level of sensitivity to detect pain pathophysiology. However, the observation that roughly one-third of our participants displayed a supposedly dysfunctional CPM response warrants further consideration. This high proportion suggests the 20% threshold may be overly conservative and limiting the sensitivity of the approach. We suggest that additional benchmark studies, providing normative data across the lifespan in pain-free individuals, are performed to ensure that the standardisation of the cuff algometer CPM paradigm also incorporates a robust standardised analysis approach.

Despite group-level differences, ICC indices of between-session test-retest reliability were *poor* for both PDT and PTT CPM-effects. These observations contrast previous studies which reported *moderate*-to-*good* ICCs for PDT CPM-effects [12,18]. Previous reports of PTT CPM-effect reliability have varied more widely, ranging from *poor* [18] to *moderate* [12]. Our Cohen’s Kappa values for responder classification were rated between *none* and *minimal* and were lower than previously described [45]. Crucially, this poor reliability cannot be attributed to fundamental measurement instability, given that the baseline PDT and PTT assessments were themselves reliable. However, CPM estimates of reliability are derived from four independent measurements, and the variability associated with each observation becomes compounded during ICC calculation [15]. While this will contribute to low reliability, it does not explain why our reliability was lower than previously reported.

ICC estimates likely also suffer from biases induced by sampling errors. ICC is the ratio of between-participant to within-participant variability [9]. Previous studies using smaller samples [4,5,12,16,18,36,37] are likely to have under-estimated between-participant variability in CPM-effects. Our bootstrapping analyses support this perspective, suggesting that ICC estimates become increasingly variable at smaller sample sizes which are prone to observing spurious and irreproducible effects [3]. These under-sampling effects may also be amplified by publication bias and file drawer practices that favour dissemination of higher reliability estimates and statistically significant findings. We suggest that wide adoption of robust, open, and transparent research practices, wherein study protocols, analyses, and dissemination plans are registered in advance, are required to ameliorate these issues [30].

Prior studies have inadequately considered the impact of ceiling effects, where participants reach the algometer’s safety limit during pressure threshold assessments. A common practice has been to assign this safety limit as the participant’s final PTT [4,12,16,45] rather than excluding the data point. This method artificially deflates the true variability in pain tolerance, leading to overestimates of PTT reliability. Consequently, it also distorts responder classifications. Our finding that 17% of individuals reached safety limits, while in line with prior reports [16], places practical limits on the applicability of PTT cuff algometry in healthy volunteers.

To classify individuals as CPM responders or non-responders, a threshold must be defined to separate them. However, normative thresholds have yet to be established, and thresholding methods proposed to date remain suboptimal. Typically, these are derived from measurement error estimates (CoV [43,44] or SEM [5,19,31,45]), but this only indicates whether observed threshold changes exceed random error. Recently, the lower 95% CI for the PDT CPM-effect of a normative sample was employed as a dysfunctional CPM threshold [34]. Whilst effective for comparing healthy samples with patient groups, in isolation this method cannot reliably indicate a response rate in healthy individuals. Both functional CPM and measurement error must both be quantified and considered to facilitate effective classification. However, measurement error estimates observed in our data are similar in magnitude to previously reported lower 95% CIs [34], with some existing error estimates exceeding this value [31]. Accordingly, where measurement error ends, and a functional CPM-effect begins, is unclear. This ambiguity highlights the inherent difficulty of imposing a binary cut-off on what is fundamentally a continuous biological process. Whilst binary categorisation is convenient and well-suited to common trial designs and statistical techniques [40], it also risks sacrificing fine-grained information that may provide mechanistic insights [49]. We suggest considering CPM readouts as continua, aligning with evolving perspectives within pain research [39], and the wider fields of neurology and psychiatry [1], where pathophysiological states are increasingly understood in this manner.

Dynamic state fluctuations also increase within-participant variance estimates considered during ICC calculation, lowering reliability estimates [9]. An individual’s emergent pain experience is tempered by competing motivational demands including, but not limited to, physiological stress, perceived threat, selective attention, prior experiences, arousal state, alertness, and circadian effects [6,24,28,42]. Pre-clinical work examining diffuse noxious inhibitory control mechanisms (DNIC), a core element of the neural circuitry proposed to underpin CPM, suggests that propriospinal activity can also influence its expression [29], in addition to the well-described descending brainstem circuitry [21,22]. However, unlike assessments made in anaesthetised animal preparations, state fluctuations in top-down control pathways occur in wakeful humans that constantly modulate CPM responses. Future longitudinal studies combining psychophysics with neuroimaging could uncover some of the mechanisms underpinning this dynamic process. [17].

Our work is not without limitations. First, our findings are specific to the young, healthy cohort studied and may not generalize to older individuals or clinical populations who often exhibit altered CPM [14]. Second, while conducting the study at a single site with a standardized protocol ensured high experimental control, our results may not capture the full variability that would arise from a multi-site study. Similarly, though the use of multiple experimenters reflects a real-world scenario, we acknowledge their contributions to the dataset were not uniform; however, this was mitigated in the crucial test-retest analysis, where data were collected by only two individuals. Finally, while computer-controlled cuff algometry is designed to be user-independent, some procedural variability, such as in cuff placement, was likely and unavoidable.

We have demonstrated that while cuff algometry produces robust group-level CPM effects, between-session reliability was poor. These findings echo growing contention regarding the clinical utility of CPM [7] including its suitability as a biomarker. Like others, we propose that state-dependent effects render single time point measurement of CPM a poor index of an individual’s overall endogenous pain control capacity [29]. We urge that the conceptualisation of CPM as a trait measure of endogenous descending control should be reconsidered in favour of a construct that reflects both static between-individual effects alongside dynamic within-individual variability.

## Data Availability

Data are not available

## Acknowledgements

Authors would like to thank to the ten experimenters who contributed to data collection alongside the authors. JLT is in receipt of a PhD studentship funded by the National Institute for Health Research (NIHR) Biomedical Research Centre at South London and Maudsley NHS Foundation Trust and King’s College London. OSK is supported by the King’s Prize Fellowship, King’s College London. TGN receives funding from the Lundbeck Foundation (R441-2023-232) and is a part of the Center for Neuroplasticity and Pain (CNAP) which is supported by the Danish National Research Foundation (DNRF121). MAH is supported by the NIHR Biomedical Research Centre and Clinical Research Facility at South London and Maudsley NHS Foundation Trust and King’s College London. KB is supported by a Medical Research Council grant (MR/W004739/1). MAH, OSK, JT and KB were also supported by the Medical Research Council (MR/N026969/1). The views expressed are those of the authors and not necessarily those of the NHS, NIHR, Medical Research Council, or Department of Health and Social Care.

