## Supplementary for "Cuff Algometry Induces Large Yet Variable Conditioned Pain Modulation Effects"

**Supplementary Information**

Joseph L. Taylor^1,2^*, Timothy Lawn^2,3^*, Olivia S. Kowalczyk^2,4^,

Thomas Graven-Nielsen^5^, Matthew A. Howard^2#^, Kirsty Bannister^6#^

^1^	Wolfson Sensory Pain and Regeneration Centre, Institute of Psychiatry, Psychology and Neuroscience, King’s College London, London, UK

^2^	Department of Neuroimaging, Institute of Psychiatry, Psychology and Neuroscience, King’s College London, London, UK

^3^	Athinoula A. Martinos Center for Biomedical Imaging, Department of Radiology, Massachusetts General Hospital and Harvard Medical School, Boston, MA, USA

^4^	Department of Imaging Neuroscience, Queen Square Institute of Neurology, University College London, UK

^5^	Center for Neuroplasticity and Pain (CNAP), Department of Health Science and Technology, Faculty of Medicine, Aalborg University, Aalborg, Denmark

^6^	Department of Life Sciences, Faculty of Natural Sciences, Imperial College London, London, UK

These authors contributed equally (joint first* and joint last^#^ authorship)

**Corresponding author**

Joseph L. Taylor

Address: L1.08, 16 De Crespigny Park, Centre for Neuroimaging Sciences, Institute of Psychiatry, Psychology and Neuroscience, King’s College London, London, SE5 8AF

ORCID ID: 0000-0003-4331-2162

| **Supplementary Table 1.**  **Missing Data for Variables Included Within Main Analyses (N = 324)** | |
| --- | --- |
| **Variable** | **Number of Missing Entries (Percentage of Total)** |
| Age | 7 (2.16) |
| Sex | 1 (0.31) |
| Baseline dominant-leg PDT | 0 (0) |
| Baseline dominant-leg PTT | 0 (0) |
| Baseline non-dominant-leg PDT | 1 (0.31) |
| Baseline non-dominant-leg PTT | 0 (0) |
| Conditioned dominant-leg PDT | 6 (1.85) |
| Conditioned dominant-leg PTT | 5 (1.54) |

Note: These numbers do not directly lead to the final sample size of 311/257 due to participants with multiple missing values.

| **Supplementary Table 2.**  **Correlation matrix of all measurements taken** | | | | | | | | |
| --- | --- | --- | --- | --- | --- | --- | --- | --- |
|  | Dominant-leg PDT | Non-dominant-leg PDT | PDT with Conditioning | CPM-effect PDT | Dominant-leg PTT | Non-dominant-leg PTT | PTT with Conditioning | CPM-effect PTT |
| Dominant-leg PDT |  |  |  |  |  |  |  |  |
| Non-dominant-leg PDT | .729^**^ (311) |  |  |  |  |  |  |  |
| PDT with Conditioning | .761^**^ (311) | .772^**^ (310) |  |  |  |  |  |  |
| CPM-effect PDT | .175^**^ (311) | .456^**^ (310) | .772^**^ (311) |  |  |  |  |  |
| Dominant-leg PTT | .571^**^ (257) | .606^**^ (256) | .568^**^ (256) | .259^**^ (256) |  |  |  |  |
| Non-dominant-leg PTT | .530^**^ (257) | .678^**^ (256) | .587^**^ (256) | .328^**^ (256) | .857^**^ (257) |  |  |  |
| PTT with Conditioning | .562^**^ (257) | .623^**^ (256) | .613^**^ (256) | .339^**^ (256) | .923^**^ (257) | .886^**^ (257) |  |  |
| CPM-effect PTT | .134^*^ (257) | .212^**^ (256) | .273^**^ (256) | .277^**^ (256) | 0.076 (257) | .311^**^ (257) | .455^**^ (257) |  |
| *Note.* Pearson correlation coefficient (n) ** = p < .01, * = p < .05. Green=PDT, Orange=PTT, Blue=PDTxPTT | | | | | | | | |

**Supplementary Figure 1. Exploration of Sex Differences within the Main Sample**

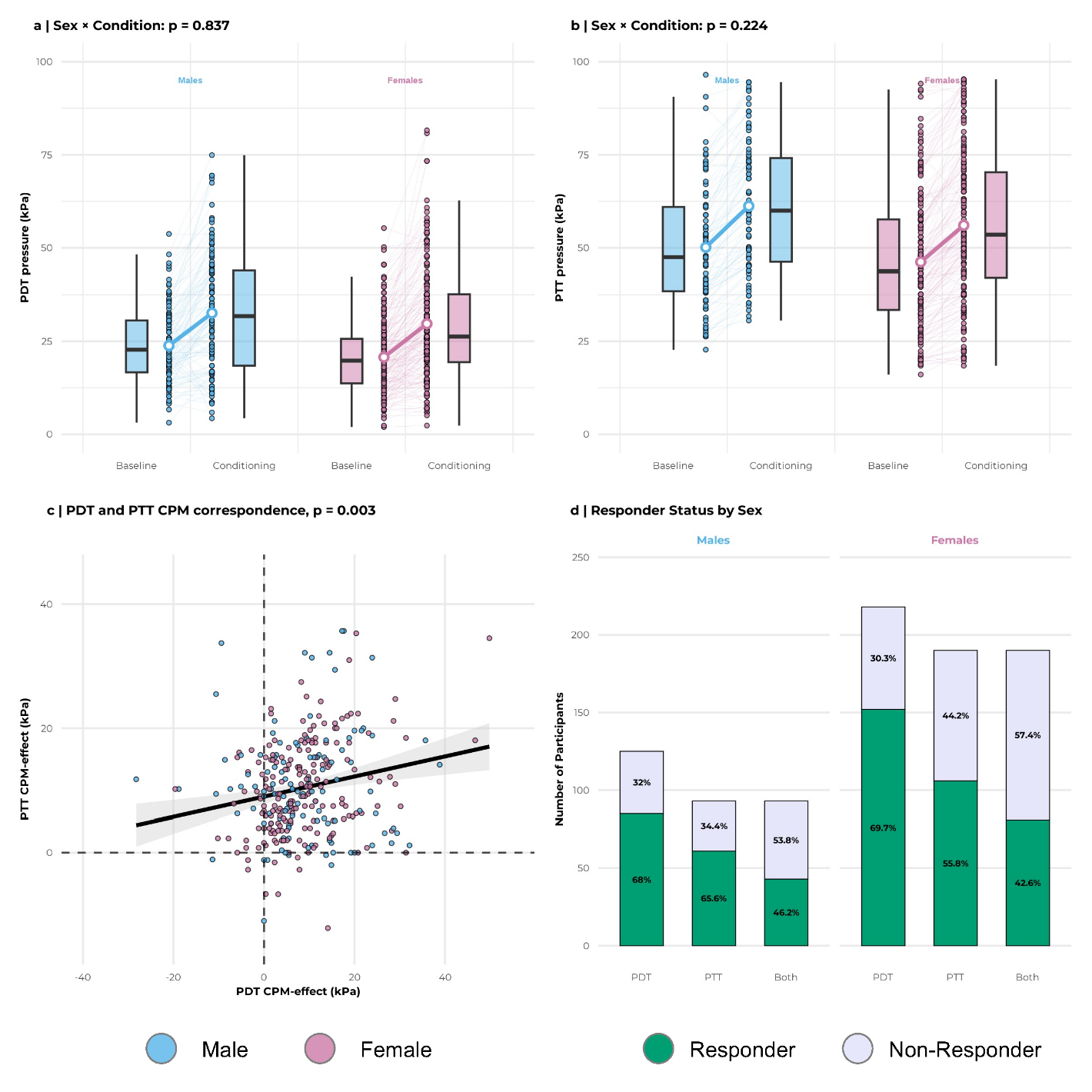

**This exploratory analysis examined sex differences in CPM effects. While absolute pain thresholds differed significantly between sexes, the magnitude of the CPM effect was equivalent for both males and females.** Linear mixed-effects models were used to analyse both Pain Detection Thresholds (PDT) and Pain Tolerance Thresholds (PTT) (a) Male participants had significantly higher PDTs than female participants overall (main effect of sex: p=0.004). For both sexes, thresholds increased significantly during conditioning (main effect of Condition: p<0.001). However, there was no significant interaction between sex and condition (p=0.84), indicating that the magnitude of the PDT CPM-effect did not differ between males and females. (b) A similar pattern was observed for PTT. Males had significantly higher pain tolerance overall (main effect of sex: p=0.004), and conditioning significantly increased thresholds for both groups (main effect of condition: p<0.001). Again, the non-significant interaction (p=0.224) demonstrates that the PTT CPM-effect was comparable across sexes. Shaded areas represent the 95% confidence interval.

**Supplementary Figure 2. Reliability of the Relative (Change from Baseline) CPM-Effect**

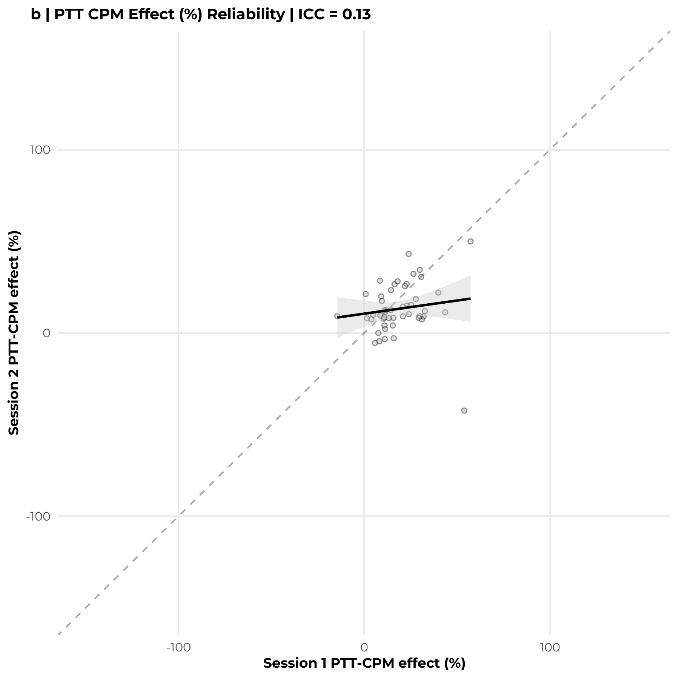

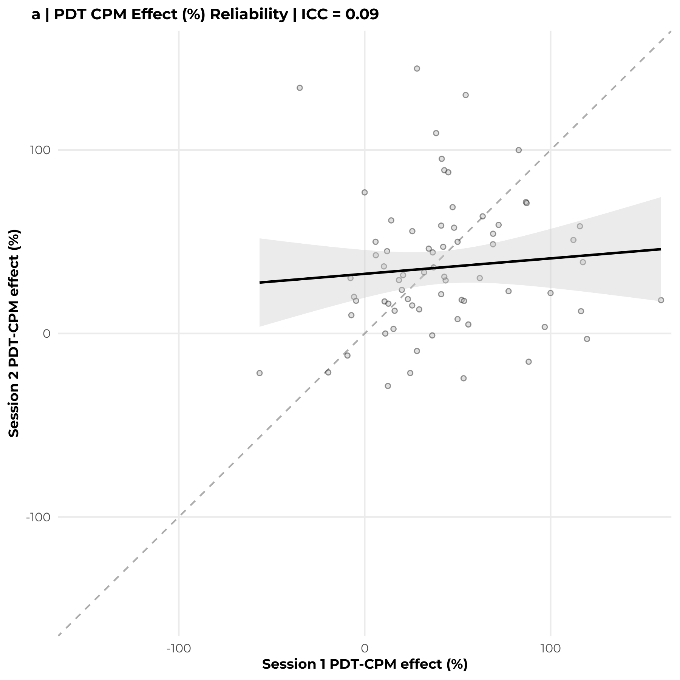

**This sensitivity analysis shows that calculating the CPM-effect as a relative (percentage) change from baseline, rather than an absolute value, does not improve its poor test-retest reliability.** The scatterplots display the relationship between Session 1 and Session 2 for the (a) Pain Detection Threshold (PDT) and (b) Pain Tolerance Threshold (PTT). Reliability remained poor for both PDT (ICC = 0.09) and PTT (ICC = 0.33). For PDT, the mean (±SD) change was 41.42% (±39.27%) in Session 1 and 36.03% (±37.50%) in Session 2. For PTT, the mean change was 16.81% (±13.65%) in Session 1 and 10.09% (±13.20%) in Session 2. One participant was excluded from the PTT analysis due to an extreme outlier generated by the percentage change calculation. Shaded areas represent the 95% confidence interval.

**Supplementary Figure 3. CPM Responder Rates and Reliability at a Range of Percentage Change Thresholds**

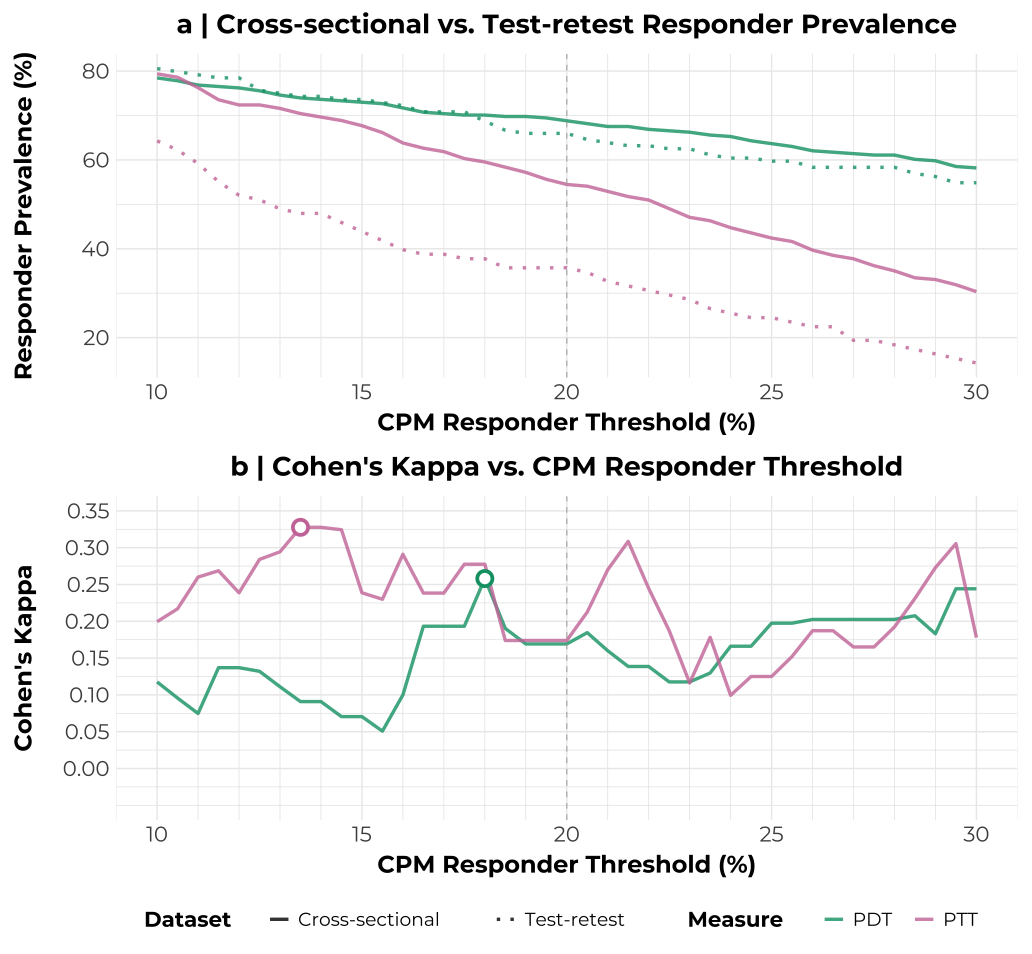

**This sensitivity analysis was performed to determine if the choice of the 20% responder threshold influenced our conclusions.** We calculated responder prevalence and test-retest reliability (Cohen's Kappa) across a range of potential thresholds from 10% to 30%. (a) As expected, the proportion of participants classified as responders steadily decreased as the threshold for classification became more stringent. (b) Test-retest reliability, assessed by Cohen's Kappa, remained consistently poor regardless of the threshold. All calculated Kappa values for both PDT and PTT fell within the 'none to minimal' agreement range. Overall, no 'optimal' threshold emerged from this analysis. This analysis demonstrates that our main finding of poor reliability is robust, rather an artifact of the specific 20% cut-off threshold used in the main text.
